# Therapeutic Prospects for Th-17 Cell Immune Storm Syndrome and Neurological Symptoms in COVID-19: Thiamine Efficacy and Safety, In-vitro Evidence and Pharmacokinetic Profile

**DOI:** 10.1101/2020.08.23.20177501

**Authors:** Vatsalya Vatsalya, Fengyuan Li, Jane Frimodig, Khushboo S. Gala, Shweta Srivastava, Maiying Kong, Vijay A Ramchandani, Wenke Feng, Xiang Zhang, Craig J McClain

## Abstract

**Introduction:** Emerging infectious diseases, especially the coronavirus disease identified in 2019 (COVID-19), can be complicated by a severe exacerbation in the Th17 cell-mediated IL-17 proinflammatory immune storm. This enhanced immune response plays a major role in mortality and morbidity, including neurological symptoms. We hypothesized that countering the cytokine storm with thiamine may have therapeutic efficacy in lowering the Th17 cell proinflammatory response. We used an *in vitro* study and corroborated those results in disease controls (DC). We developed an effective dose range and model for key pharmacokinetic measures with the potential of targeting the cytokine storm and neurological symptoms of COVID-19.

**Study Participants and Methods:** We investigated the effect of a three-week 200 mg dose of thiamine in lowering the Th17 response in sixteen DC (proinflammatory origin due to heavy alcohol drinking) patients; and eight healthy control/volunteers (HV) as a pilot clinical-translational investigation. To further investigate, we performed an *in vitro* study evaluating the effectiveness of thiamine treatment in lowering the Th17 proinflammatory response in a mouse macrophage cell line (RAW264.7) treated with ethanol. In this *in vitro* study, 100 mg/day equivalent (0.01 µg/ml) thiamine was used. Based on recent publications, we compared the results of the IL-17 response from our clinical and *in vitro* study to those found in other proinflammatory disease conditions (metabolic conditions, septic shock, viral infections and COVID-19), including symptoms, and dose ranges of effective and safe administration of thiamine. We developed a dose range and pharmacokinetic profile for thiamine as a novel intervention strategy in COVID-19 to alleviate the effects of the cytokine storm and neurological symptoms.

**Results:** The DC group showed significantly elevated proinflammatory cytokines compared to HV. Three-week of 200 mg daily thiamine treatment significantly lowered the baseline IL-17 levels while increased IL-22 levels (anti-inflammatory response). This was validated by an *in vitro* macrophage response using a lower thiamine dose equivalent (100 mg), which resulted in attenuation of IL-17 and elevation of IL-22 at the mRNA level compared to the ethanol-only treated group. In humans, a range of 79-474 mg daily of thiamine was estimated to be effective and safe as an intervention for the COVID-19 cytokine storm. A literature review showed that several neurological symptoms of COVID-19 (which exist in 45.5% of the severe cases) occur in other viral infections and neuroinflammatory states that may also respond to thiamine treatment.

**Discussion:** The Th17 mediated IL-17 proinflammatory response can potentially be attenuated by thiamine. Thiamine, a very safe drug even at very high doses, could be repurposed for treating the cytokine/immune storm of COVID-19 and the subsequent neurological symptoms observed in COVID-19 patients. Further studies using thiamine as an interventional/prevention strategy in severe COVID-19 patients could identify its precise anti-inflammatory role.

## Introduction

Viral diseases and wide-spread outbreaks have adverse health-related consequences worldwide. Emerging infectious diseases (EID) include viral pathogens that have shown higher incidence of human infection in the past several decades and raise concerns regarding increased ongoing/future prevalence^1^. Coronavirus is recognized as an EID that has become a challenging and aggressive infection with high morbidity and mortality in humans^2^. SARS-CoV-2 (severe acute respiratory syndrome coronavirus 2; causes coronavirus disease [COVID-19]) was identified in 2019, has become a pandemic, and is a priority healthcare concern in the year 2020^3^.

In viral infections, tissue inflammation is driven by multiple proinflammatory and immunoregulatory signals^4,5^. The pathological progression of COVID-19 has multiple clinical stages and may present with the cytokine storm syndrome^6^ and immunosuppression^7^. Interleukin-17 (IL-17) is a cytokine^8^ that is often involved in a proinflammatory response in the cytokine storm of viral infections^9–11^. It can also promote respiratory viral infections^12^, tissue pathology^13–15^, and neurological manifestations^16^. Th17 cells also produce Interleukin-22 (IL-22), which plays a protective/anti-inflammatory role, and it is dysregulated in several proinflammatory conditions^17^. Thus, a therapy that could alleviate the Th17 mediated pro-inflammatory response^18^ might be effective in attenuating the cytokine storm observed in COVID-19 patients.

Thiamine, a vitamin and dietary supplement,^19^ has anti-oxidant properties^19,20^. High levels of cytokines (for example, IL-1β and IL-6) may occur in thiamine deficient subjects and can be associated with oxidative stress and inflammation^21,22^. Importantly, thiamine administration could inhibit production of these cytokines, increase anti-inflammatory activity^23,24^, and potentially alleviate neuroinflammatory symptoms of viral origin^25,26^.

We tested the efficacy of a three-week thiamine treatment in modulating the Th17 proinflammatory response in a human disease control model of conditions associated with inflammation. To validate the effectiveness of thiamine in treating the proinflammatory response from the human study, we conducted an *in vitro* experiment to test the effects of thiamine treatment in alleviating ethanol mediated immune dysregulation in a mouse macrophage cell line, RAW264.7. We investigated the Th17 cells proinflammatory cytokine response (namely IL-17) in both healthy controls and individuals with high inflammatory response. This was done to estimate the effects of various doses of thiamine that have shown efficacy in alleviating the Th17 associated cytokine response. We assessed the pharmacokinetics of the oral thiamine dosing. Lastly, we also examined the neurological symptoms of COVID-19 that could possibly be treated with thiamine.

## Study Participants and Methods

### Study Participants

This investigation was approved under two large clinical investigations that were conducted at the University of Louisville (NCT#01809132, HV cohort), and the National Institute on Alcohol Abuse and Alcoholism (NIAAA) (NCT#00106106, DC cohort) at the National Institutes of Health (NIH), Bethesda MD. The studies were approved by the NIH Institutional Review Board (IRB) committee and the UofL IRB (IRB # 12.0427). All DC (disease controls as termed in this study, who were alcohol use disorder [AUD]) patients received acamprosate (or placebo) as part of a larger addiction intervention investigation. All of these patients also received thiamine as part of the medical management, which is the primary aim of this study. Sixteen age- and sex- matched male and female alcohol use disorder (AUD) patients (Termed as disease controls [DC] in this investigation) between 21-65 yrs. of age with both present and past heavy drinking profile participated as the DC, who were diagnosed with AUD based on DSM-IV TR criteria. They received daily doses of open label thiamine (100 mg twice daily = 200 mg per day)^27^ for 3-weeks after completion of the consenting process. DC patients also received standard of care inpatient medical management, including counseling. Detailed information on subject recruitment and management can be obtained from several of our previous publications^28–31^. We also included eight healthy controls in this study for comparison with DC. Demographic data were collected from all the participants. Baseline (HV and DC) and post-treatment (DC only) blood samples (after the completion of 3-weeks of thiamine dosing) were collected, processed (for plasma extraction) and frozen at −80°C. They were subsequently thawed and assayed. *Laboratory Assays and Therapeutic Model on Th17 Inflammation Axis*

#### (1) Cytokine assays

Plasma levels of proinflammatory cytokines, IL-1β, IL-6, and IL-10 were obtained by multianalyte chemiluminescent detection using Multiplex kits (Millipore, Billerica, MA) on the Luminex platform (Luminex, Austin, TX), according to manufacturers’ instructions.

#### (2) Analysis of IL-17 and IL-22 in a set of AUD patients for designing proof-of-concept experimental model

We performed analyses for IL-17 and IL-22 on human plasma samples to estimate the Th17 inflammatory response, with the goal of developing an *in vitro* mechanistic experimental model to test the efficacy of thiamine. The plasma levels of IL-17 and IL-22 in eight healthy volunteers were also included in this study for comparison. IL-17 and IL-22 were detected in plasma using Human IL-17A (now called IL-17) High Sensitivity ELISA Kits (BMS2017HS, Invitrogen) and Human IL-22 ELISA Kits (BMS2047, Invitrogen) per the manufacturer’s instructions. Results were read on a Spectra Max Plus 384 plate reader and modeled using their SoftMax Pro software (Molecular Devices, San Jose, CA).

#### (3) Cell culture

RAW 264.7 cells (mouse macrophage cell line) were cultured in Dulbecco’s modified Eagle’s medium (DMEM, Invitrogen), supplemented with 10% fetal bovine serum (FBS) and 1% penicillin/streptomycin. Cells were seeded in a 24-well culture plate and maintained at 37 °C in a 5% CO_2_ incubator for 3 days. The 0.02 µg/mL treatment dose was equivalent to the 200 mg/day thiamine dose (approximate blood AUC = 204 nmol/L^32^) given to the patients. Cells were then treated with thiamine (Vit B1 [V_B1_] as shown in the Figure 2) at a concentration of (0.01 µg/mL) for 2 hours (in a preventive paradigm), followed by 80 mM ethanol treatment for 22 hours, for a total of 24 hours of treatment to determine the minimum effective level of thiamine to reduce the Th17 response. Cells were then washed with PBS and collected with Trizol reagent for the isolation of RNA. RNA samples were reverse transcribed to cDNA and used for qRT PCR analysis of cytokine expression (IL-17, IL-22). Cell viability was not affected by thiamine or EtOH treatment at the doses used in the experiments.

**Figure 1:**
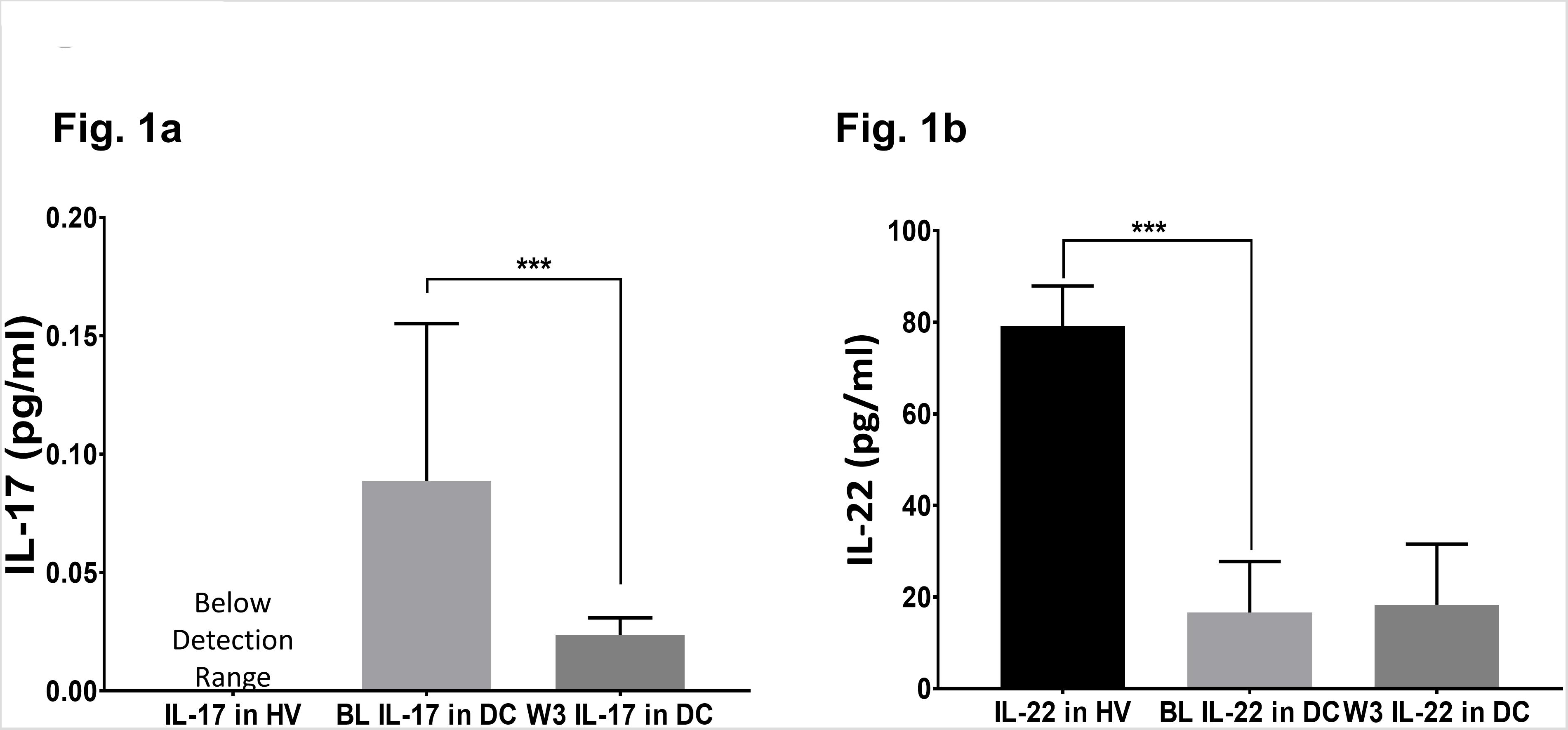
Efficacy of thiamine treatment on Th17 cell derived response for IL-17, and IL-22 cytokines. Levels of IL-17 and IL-22 in healthy volunteers (HV) at baseline; and Disease Controls (DC or [alcohol use disorder, AUD]) patients exhibiting a proinflammatory response (n = 16) at baseline; and anti-inflammatory normalization of cytokines tested after the completion of three-week (W3) thiamine treatment. A drop in IL-17 coupled with a mild increase in IL-22 at W3 was observed compared to the baseline levels. BL: baseline, W3: three-week of thiamine treatment. Data are presented as M±SD. Statistical significance was set as p < 0.05.

**Figure 2:**
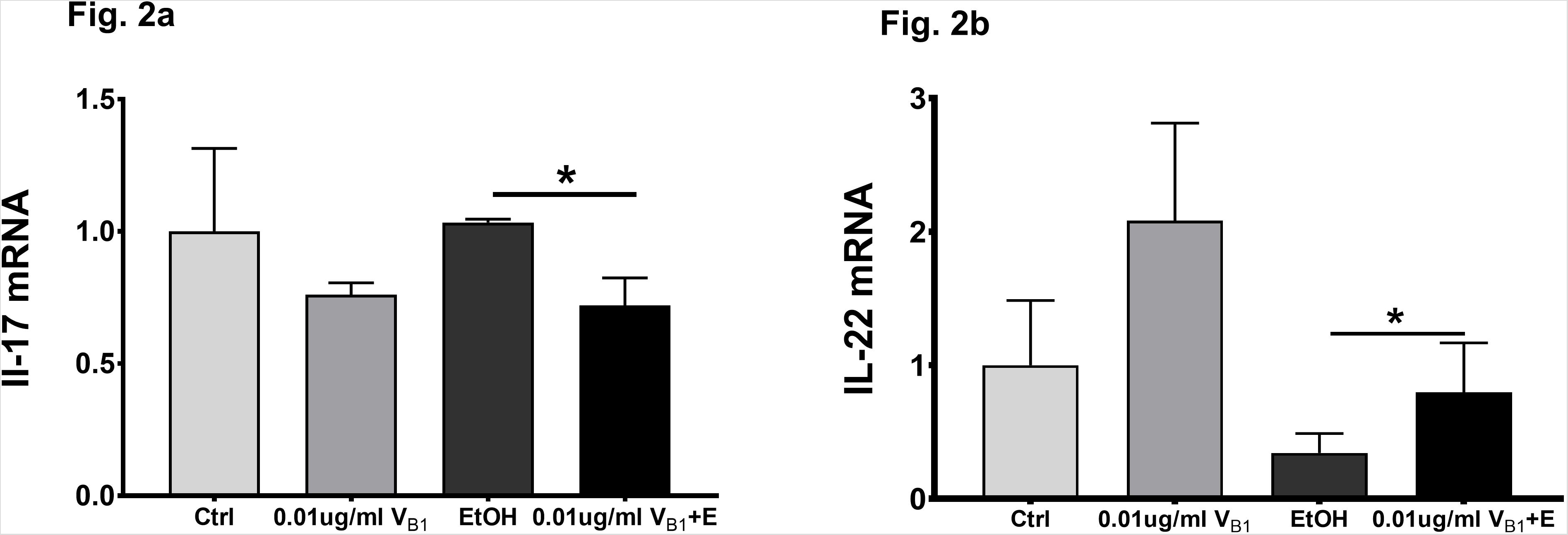
mRNA expression of IL-17 and IL-22 in an *in vitro* model of the mouse macrophage cell line using a potent proinflammatory agent (alcohol), and preventative administration of thiamine. Fig. 2a: Significant lowering was observed in IL-17 mRNA expression in “EtOH + V_B1_” (Bar 4) group compared to the alcohol-treated group (Bar 3). Fig. 2b: Significant elevation was observed in IL-22 mRNA expression in “EtOH + V_B1_” (Bar 4) group compared to the alcohol-treated group (Bar 3). Controls (both Fig. 2a and 2b) show normal anti-inflammatory effects of thiamine on IL-17 and IL-22 response with thiamine administration. Ctrl: Non-treated controls. V_B1_ 0.01 ug/ml. EtOH: alcohol treated. V_B1_+E: thiamine and alcohol treated. Data are presented as M±SD. Statistical significance was set as p < 0.05.

#### (4) RNA isolation and real-time RT-PCR

Total RNA was extracted from the cells using Trizol reagent (500 µl/well) according to manufacturer’s instruction (Life Technologies, Carlsbad, CA) and reverse-transcribed using cDNA Supermix (QuantaBio, Beverly, MA). Quantitative real-time PCR was performed on an ABI 7500 real-time PCR thermocycler and SYBR green PCR Master Mix (Applied Biosystems, Foster City, CA) was used for quantitative real-time PCR analysis. The relative quantities of target transcripts were calculated from duplicate samples after normalization of the data against the housekeeping gene, mouse 18S. Relative mRNA expression was calculated using comparative Ct method. The following primer pairs were used:

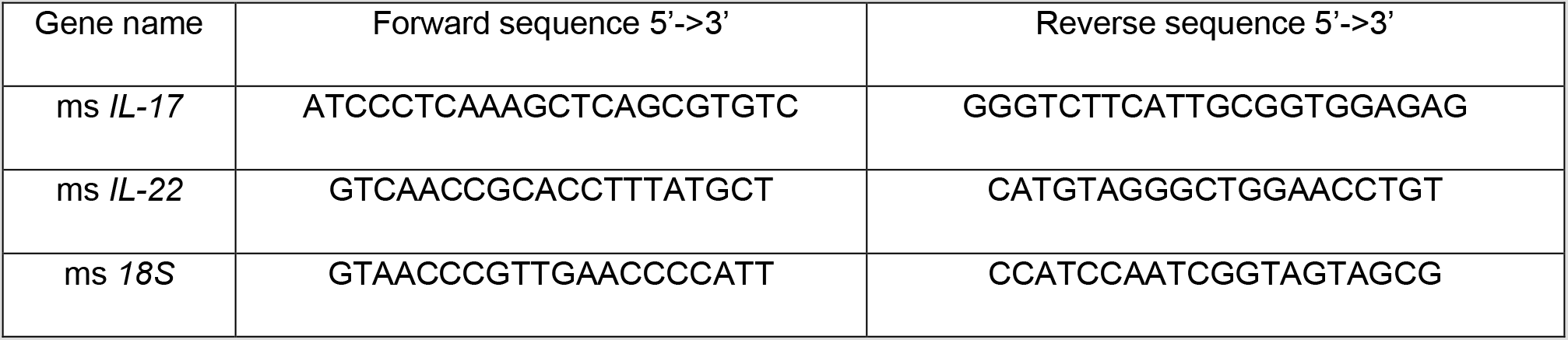

### Development of the Pharmacokinetic Model for Dose Titration of Thiamine

We used dosing guidelines for thiamine as mentioned at the Medline Plus (https://medlineplus.gov/druginfo/natural/965.html#Safety, last reviewed as of August 5, 2020), and from peer reviewed publications from PubMed (https://pubmed.ncbi.nlm.nih.gov/; [searched and collected until August 5, 2020]). We used available dosing guidelines from Medline Plus for healthy individuals both for dietary supplementation and vitamin deficiency status. We also reviewed and incorporated thiamine dose levels (lower and higher range) from other disease conditions; namely metabolic conditions^33,34^, septic shock^35,36^, viral diseases^37–39^ and Leigh’s disease^40^ (Medline Plus: Thiamine). We also included the recorded thiamine dose levels from the DC group (AUD with Wernicke Korsakoff Syndrome, WKS^41^; from our clinical study) as one of the pro-inflammatory conditions.

We compared the reference range of levels of the Th17 cytokine (IL-17) response in disease/health conditions in humans as published in the recent findings concerning COVID-19’s cytokine storm data^42–44^. A Th17 proinflammatory response for the potential range of IL-17 levels was also developed for healthy volunteers (HV, from our study cohort), metabolic conditions^45,46^, DC (or alcohol use disorder patients from our study cohort), septic shock^47,48^, and viral infections^12,49^. IL-17 data on severe COVID-19 patients (as mentioned above) were collected from the recently peer-reviewed published articles found in PubMed (searched until August 5, 2020). Doses administered to our DC study cohort, and data from healthy individuals (HV) were also used in the development of the dose profile. All these data were incorporated in the predictive regression model for identifying a tentative effective dose range of thiamine (Figure 3).

The pharmacokinetic response of thiamine was calculated at both the low and high ends of the dose range described above. The area under the curve and maximum concentration (C_max_) were established for both blood and plasma for a 10-hour trajectory (Figure 4) using the indices of thiamine’s *in vivo* blood pharmacokinetics^32^. For the derived 79 mg thiamine dosing (low end), the slopes used to identify AUC in blood were 2.14, and 1.76 in plasma. For the 474 mg thiamine dosing (upper end), the slope used to derive AUC in blood was 1.02, while in plasma it was 1.09. Similarly, for the 79 mg thiamine dosing, the slope used to derive C_max_ in blood was 0.40, and in plasma it was 0.39. For the 474 mg thiamine dose, the slope used to derive AUC in blood was 0.14, and in plasma it was 0.18.

*Statistical Analysis*

Data are expressed as Mean ± standard deviation (M±SD) in Table 1 as well as in Figures 1 and 2. Two-sided Student’s t-test was used to examine the difference between disease controls and healthy volunteers at baseline (see Suppl. Table 1 and Fig. 1), and two-sided Student’s paired t-test was used to examine the changes at baseline versus 3 weeks for disease controls (Figure 1).

**Table 1.**
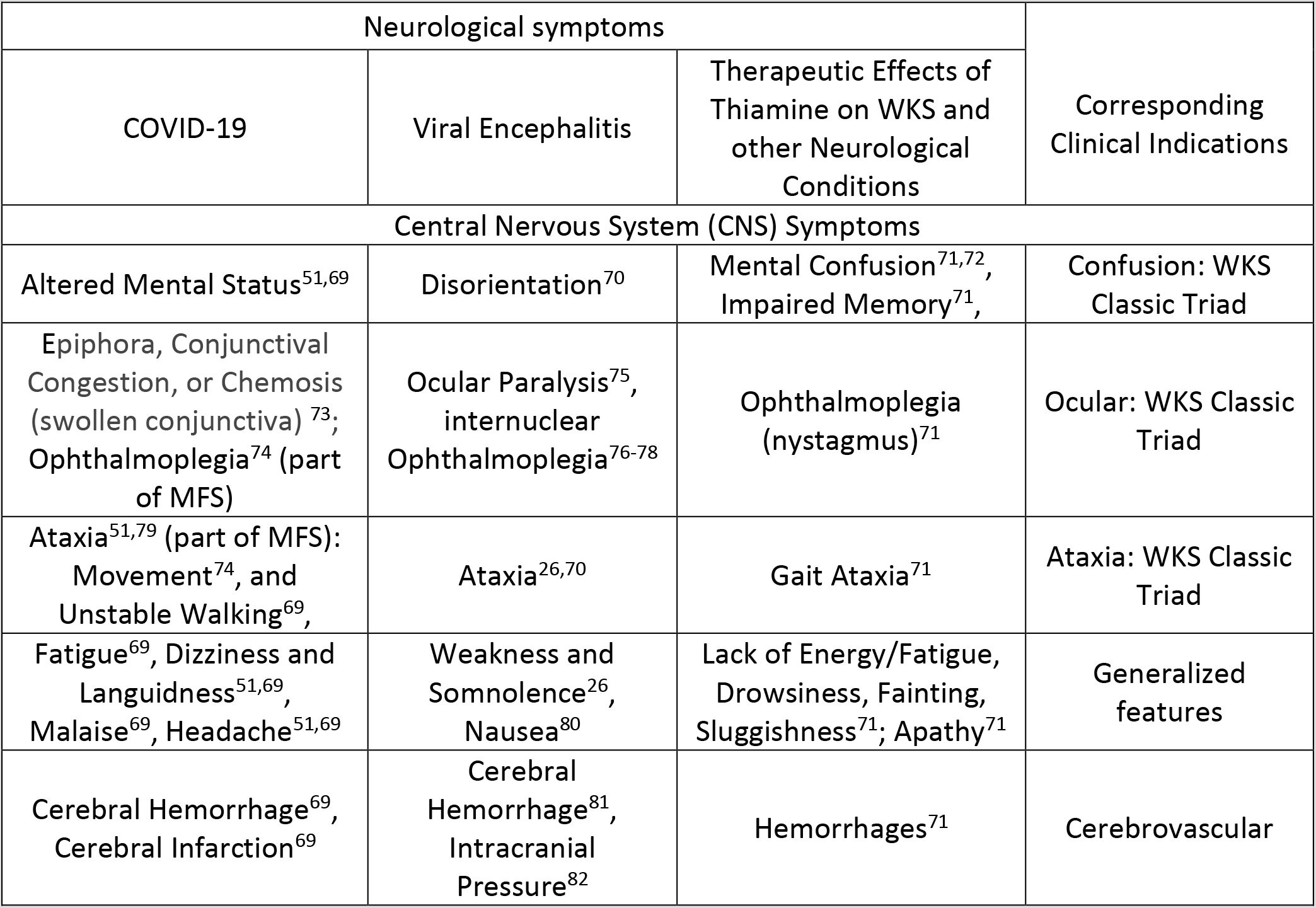
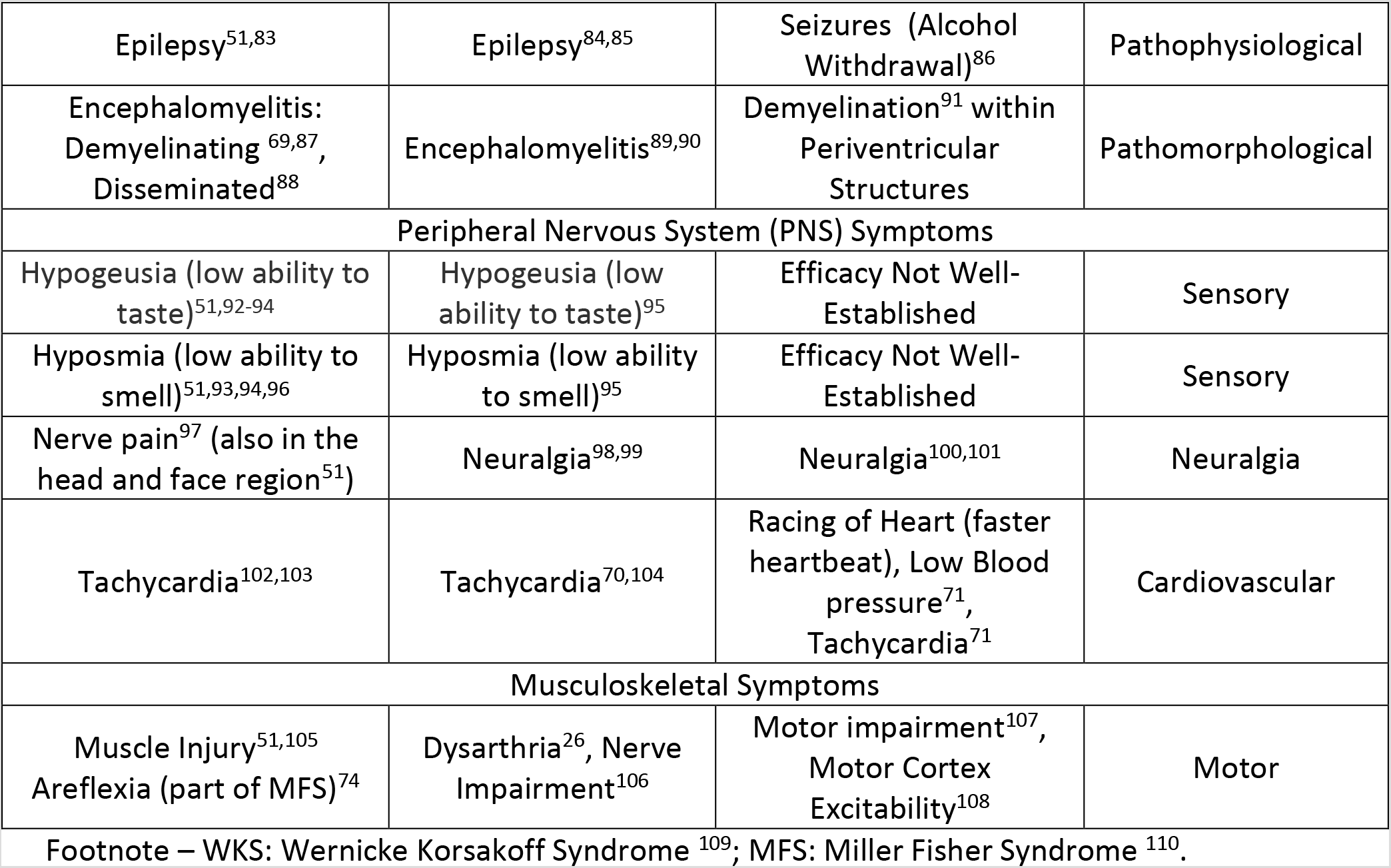
Neurological symptoms in COVID-19, Viral Encephalitis (Grouped/assorted by the neurological spectrum observed in COVID-19), showing their proximity in presentation. Therapeutic effects of thiamine on the corresponding neurological symptoms of pro-inflammatory origin that are also observed in viral infection.

| Neurological symptoms |  |  | Corresponding Clinical Indications |
| --- | --- | --- | --- |
| COVID-19 | Viral Encephalitis | Therapeutic Effects of Thiamine on WKS and other Neurological Conditions |  |
| Central Nervous System (CNS) Symptoms |  |  |  |
| Altered Mental Status<sup>51,69</sup> | Disorientation<sup>70</sup> | Mental Confusion<sup>71,72</sup>, Impaired Memory<sup>71</sup>, | Confusion: WKS Classic Triad |
| Epiphora, Conjunctival Congestion, or Chemosis (swollen conjunctiva) <sup>73</sup>; Ophthalmoplegia<sup>74</sup> (part of MFS) | Ocular Paralysis<sup>75</sup>, internuclear Ophthalmoplegia<sup>76-78</sup> | Ophthalmoplegia (nystagmus)<sup>71</sup> | Ocular: WKS Classic Triad |
| Ataxia<sup>51,79</sup> (part of MFS): Movement<sup>74</sup>, and Unstable Walking<sup>69</sup>, | Ataxia<sup>26,70</sup> | Gait Ataxia<sup>71</sup> | Ataxia: WKS Classic Triad |
| Fatigue<sup>69</sup>, Dizziness and Languidness<sup>51,69</sup>, Malaise<sup>69</sup>, Headache<sup>51,69</sup> | Weakness and Somnolence<sup>26</sup>, Nausea<sup>80</sup> | Lack of Energy/Fatigue, Drowsiness, Fainting, Sluggishness<sup>71</sup>; Apathy<sup>71</sup> | Generalized features |
| Cerebral Hemorrhage<sup>69</sup>, Cerebral Infarction<sup>69</sup> | Cerebral Hemorrhage<sup>81</sup>, Intracranial Pressure<sup>82</sup> | Hemorrhages<sup>71</sup> | Cerebrovascular |
| Epilepsy<sup>51,83</sup> | Epilepsy<sup>84,85</sup> | Seizures (Alcohol Withdrawal)<sup>86</sup> | Pathophysiological |
| Encephalomyelitis: Demyelinating<sup>69,87</sup>, Disseminated<sup>88</sup> | Encephalomyelitis<sup>89,90</sup> | Demyelination<sup>91</sup> within Periventricular Structures | Pathomorphological |
| Peripheral Nervous System (PNS) Symptoms |  |  |  |
| Hypogeusia (low ability to taste)<sup>51,92-94</sup> | Hypogeusia (low ability to taste)<sup>95</sup> | Efficacy Not Well-Established | Sensory |
| Hyposmia (low ability to smell)<sup>51,93,94,96</sup> | Hyposmia (low ability to smell)<sup>95</sup> | Efficacy Not Well-Established | Sensory |
| Nerve pain<sup>97</sup> (also in the head and face region<sup>51</sup>) | Neuralgia<sup>98,99</sup> | Neuralgia<sup>100,101</sup> | Neuralgia |
| Tachycardia<sup>102,103</sup> | Tachycardia<sup>70,104</sup> | Racing of Heart (faster heartbeat), Low Blood pressure<sup>71</sup>, Tachycardia<sup>71</sup> | Cardiovascular |
| Musculoskeletal Symptoms |  |  |  |
| Muscle Injury<sup>51,105</sup><br>Areflexia (part of MFS)<sup>74</sup> | Dysarthria<sup>26</sup>, Nerve Impairment<sup>106</sup> | Motor impairment<sup>107</sup>, Motor Cortex Excitability<sup>108</sup> | Motor |

Footnote – WKS: Wernicke Korsakoff Syndrome <sup>109</sup>; MFS: Miller Fisher Syndrome <sup>110</sup>.

*Post-hoc* one-sided t-tests were performed for the IL-17 and IL-22 mRNA expression analyses for the RAW 264.7 cells testing (Figure 2). A pharmacokinetic model for anticipated therapeutic dosing range of thiamine based on IL-17 ranges in different inflammatory conditions was constructed using predictive regression computation (Figure 3). Factorial between-group ANOVA was used to evaluate demographic and cytokine profiles (Table 1). Statistical significance was established at *p* < 0.05. SPSS 26.0 (IBM Chicago, IL) and Microsoft Excel 365 (MS Corp, Redmond WA), statistical software R (https://www.r-project.org/), and Prism GraphPad (GraphPad Software, San Diego CA) were used for statistical analysis, data computation, and plotting the figures.

**Figure 3:**
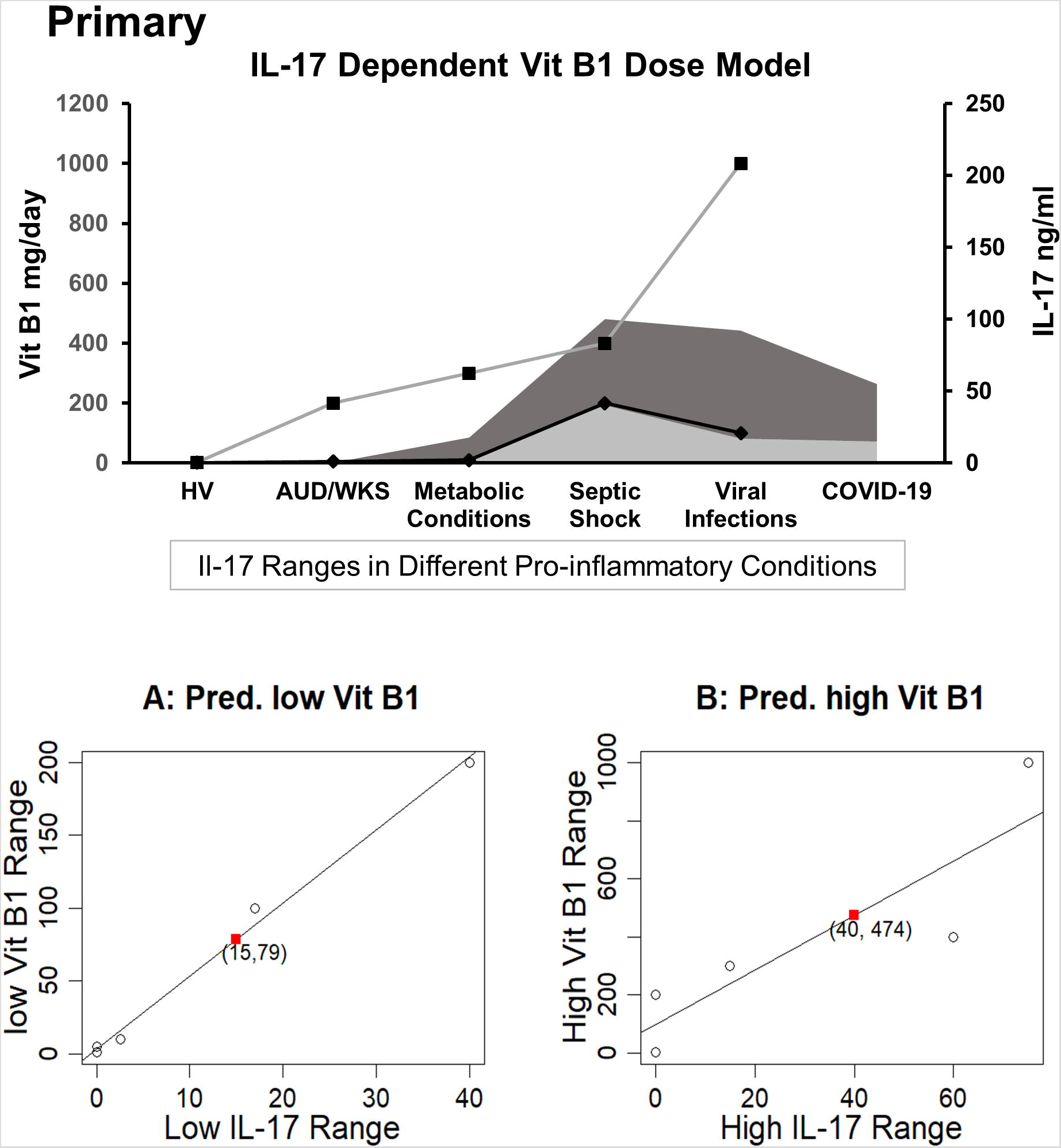
PRIMARY: A dose titration by disease and proinflammatory Th17 status model of thiamine administration with parallel representation of Th17 proinflammatory response in various groups including healthy volunteers and disease groups with pro-inflammatory response. Fig. 3A: Linear regression model is predictive for the relation between the low dose thiamine (Vit B1) range versus low IL-17 ranges based on known observed pairs from different patients; lower range of vit B1 dose (79 mg/daily) corresponding to lower range of IL-17 (15 ng/ml) in the COVID-19 patients. Dark grey shade depicts higher IL-17 response and lighter grey shade shows lower IL-17 response in various proinflammatory conditions. Fig. 3B: Linear regression shows the predictive model for the relation between a higher range of Vit B1 dose versus a higher range of IL-17 levels, derived from the known observed pairs from different patients; higher range of Vit B1 dose (474 mg/daily) corresponding to higher range of IL-17 (40 ng/ml) reported in COVID-19 patients. High Range Thiamine Dose: Left Y-axis (primary). Low Range Thiamine Dose, Low Range IL-17 levels, and High range IL-17 levels: Right Y-axis (Secondary).

### Neurological Assessments

We conducted a review on the neurological presentation in COVID-19 and other relevant viral occurrences of encephalitis (Table 1). We identified and tabulated the neurological symptoms of COVID-19 and viral encephalitis from the recently published findings. We also described the neurological symptoms, that are generally treated effectively with thiamine (Table 1). We used PubMed and Medline Plus for disease references (searched until August 5, 2020).

## Results

### Demographics and Candidate Proinflammatory Cytokine Profile

DC group individuals in this study had significantly higher age than the healthy controls (HV) (Suppl Table 1). However, there was no significant difference in the mean BMIs between the two groups, and the sex-distribution was also similar between the two groups. Both IL-6 (∼6 fold) and IL-1β (∼3 fold) cytokines were significantly higher in the DC group (AUD patients) compared to the healthy controls/volunteers (HV) group. IL-10 was also numerically higher in the DC group.

### Clinical Findings on the Immune Response of Th17 derived IL-17 and IL-22 Axis Response, and Thiamine Efficacy and Safety

To develop a model for thiamine effects on inflammation, we assessed IL-17 and IL-22 cytokine expression (showing proinflammatory and anti-inflammatory effects, respectively). Both cytokines are produced by the Th17 cells^50^. IL-17 concentrations were below the level of detection in the HV group, but were elevated in the DC group. An approximate four-fold decrease was observed in the IL-17 concentration levels (0.09 pg/mL to 0.023 pg/mL) with a treatment dose of 200 mg thiamine daily (Estimated AUC = 204 nmol/L × hour approximately in the 10-hr. window) by the end of week 3. IL-22 was significantly decreased in DC group compared to healthy volunteers and thiamine therapy did not significantly improve levels in treated DC group individuals. No patients reported any kind of drug related adverse events; therefore, the safety profile of thiamine administration was excellent at 200 mg daily in this small pilot group.

### Experimental Model for Treatment Efficacy of Thiamine on IL-17 and IL-22 Activity

Using these clinical findings of the treatment efficacy of thiamine, we designed an *in vitro* experiment using mouse macrophage RAW264.7 cells to validate the effects of thiamine on alcohol-induced IL-17 and IL-22 expression. The results showed that a low dose of thiamine (V_B1_) decreased IL-17 expression in the absence (by 20% approximately) and/or presence of alcohol treatment (by 25% roughly) (Figure 2a). Importantly, IL-22 expression was upregulated by thiamine in both the control and ethanol-treated cells. This response in IL-22 expression was moderately reduced by alcohol but rescued by the thiamine (V_B1_) treatment (Figure 2b). Half of the treatment dose (0.02 µg/mL) that was found to be effective in humans was beneficial as a preventative dose (0.01 µg/mL) of thiamine in this *in vitro* model.

### IL-17 Dependent Dose and Pharmacokinetic Model of Thiamine

An IL-17 response dependent dose and pharmacokinetic model of thiamine administration was developed, based on responses from the pro-inflammatory disease cohorts and the corresponding thiamine dosing (controlled for by the corresponding values in healthy volunteers, as a point of reference). This model supported a tentative range of thiamine dosing for COVID-19 (Figures 3A and 3B), since the IL-17 levels are much higher in COVID-19 than in the reports from many other proinflammatory disease conditions. Using regression analysis, a range of 79 mg/day (lower end of dose range) - 474 mg/day (higher end of dose range) for thiamine administration was found to correspond to a range of 15-40 ng/mL level of IL-17 used *in vitro*.

The pharmacokinetic parameters were: Area Under the Curve (AUC), and Maximum (or peak) Concentration of a drug (C_max_). These gave a very close estimation of the specific oral thiamine dose (Figure 4). As expected, plasma values were higher for both AUC and C_max_ at higher doses and were lower at lower doses (Suppl Table 2).

**Figure 4:**
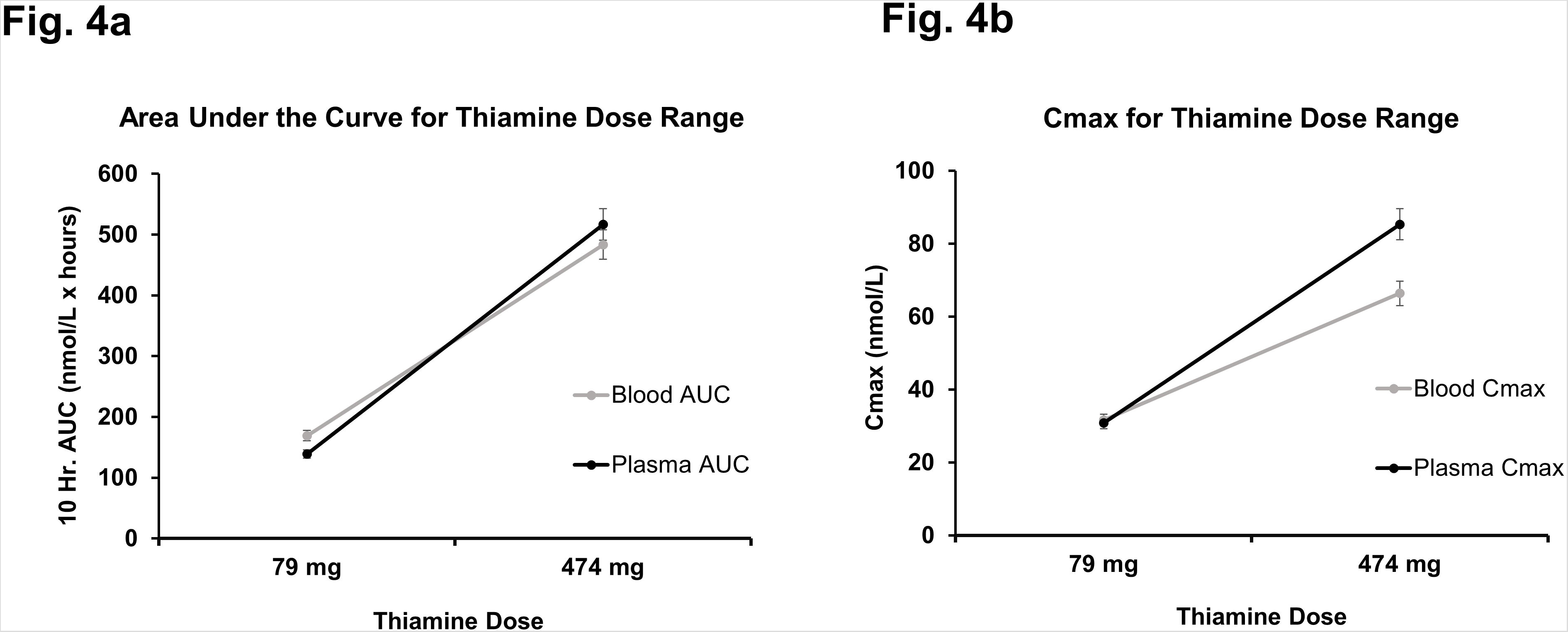
Pharmacokinetics parameters of oral thiamine over 10 hr. (Projected) in whole blood and plasma. Fig. 4a: Predicted course of Area Under the Curve (AUC) for low values (79 mg/day) and high values (474 mg/day) of the range of oral thiamine dose in blood and plasma over the 10 hours. Fig. 4b: Predicted maximum concentration of low values (79 mg/day) and high values (474 mg/day) of the range of oral thiamine dose in blood and plasma over the first 10 hours. Errors bars show a 5% variability in the pharmacokinetic measures at each dose.

### Assessment of Neurological Presentation of COVID-19, Viral Encephalitis and Therapeutic Efficacy of Thiamine Administration

We tabulated the neurological symptoms from the recently published findings on COVID-19 (Table 1). Severely ill COVID-19 patients presented with various neurologic symptoms that could be grouped together as: acute cerebrovascular disease; altered mental status; and musculoskeletal symptoms^51^. We also included the neurological presentation commonly observed in viral encephalitis of non-COVID-19 origin, grouped corresponding to the presentation of the symptoms of COVID-19.

Lastly, we also tabulated the neurological symptoms that are commonly treated with thiamine. Several neurological symptoms of COVID-19 and viral encephalitis corresponded well with the neurological spectrum that is known to be managed effectively with thiamine.

## Discussion

We evaluated individuals with significantly altered IL-17 and IL-22 responses associated with Th17 cells and found a significant role for a 3-week 200 mg/daily thiamine treatment regimen in improving the Th17 response in the AUD disease control patient cohort whose members exhibited a high pro-inflammatory status at baseline. SARS CoV-2 viral challenge causes induction of IL-6 leading to altered Th17 responses^52^. IL-17 synthesized by Th17 cells can markedly stimulate neutrophil chemotaxis and may lead to a skewed Th2 immune response^53,54^. Results from our AUD control group show the increases in candidate proinflammatory (IL-1β and IL-17), and anti-inflammatory (IL-6, IL-10 and IL-22) cytokines. Changes in IL-17 and IL-22 with alcohol abuse (pro-inflammatory), and thiamine as an anti-inflammatory therapy in our experiment provided potential proof of concept. IL-1β is a key cytokine initiating Th17 cells to synthesize IL-17^55^, whereas IL-10 suppresses Th17 proinflammatory cytokine production^56^. High IL-6 levels also are associated with Th17 cell proinflammatory activity^52^. Thus, under inflammatory conditions, there is a complex interaction of proinflammatory/anti-inflammatory responses from Th17 cells. An intervention or prevention that could attenuate the Th17 proinflammatory activity could help ameliorate the consequences of a cytokine storm.

Thiamine deficiency has been reported to promote a proinflammatory response in Th1 and Th17 cells^57^. To examine the role of thiamine in treating inflammation, *in vitro* testing was used to mechanistically examine the clinical outcomes. Our *in vitro* model showed that thiamine could effectively lower IL-17 and increase IL-22 mRNA expression in macrophages. Both our clinical data, and the *in vitro* studies suggest that thiamine could play a potential role in attenuating the cytokine storm in patients who have a strong proinflammatory response.

A study using a mouse model showed that IL-17 augments respiratory syncytial virus (RSV)-induced lung inflammation^58^. In that study, immunodepletion of IL-17 before viral infection resulted in diminished RSV-driven mucous cell hyperplasia and airspace enlargement, suggesting IL-17 as a potential therapeutic target. Proinflammatory IL-17 production could also initiate pulmonary eosinophilic response, by promoting proliferation of eosinophils in the bone marrow, followed by recruitment and extravasation into the lungs^59^. These recent findings suggest that suppression of IL-17 may be vital to managing viral infections, including COVID-19 and their harmful consequences.

We derived an effective dose range of thiamine that could be administered for alleviating the Th17 cell proinflammatory response by using the IL-17 concentrations that we obtained from our AUD patients (termed as DC), levels found in the literature, and levels in the healthy control (HV) group. Thiamine has been administered as a treatment in other viral infections^60,61^, and has proven effective for some inflammatory conditions and symptoms^62,63^. A well-structured treatment profile of thiamine based on the results of proof of concept *in vitro* experiments, and analyses of proinflammatory response-relevant disease conditions support the potential efficacy of thiamine in ameliorating the proinflammatory Th17 response in severe COVID-19 patients. Use of preventive as well as interventional dosages show potential in the management of COVID-19. Thiamine C_trough_ is reached generally in 10-12 hours; thus, the total dose prescribed can be divided into two doses per day. This may lead to fewer AEs or other side effects.

One recent report identified that 36.4% of the COVID-19 diagnosed patients have neurologic symptoms, and this proportion was higher (45.5%) among those COVID-19 patients with more severe symptoms^51^. Patients with other viral diseases have also shown clinical symptoms of beriberi^64^, or Wernicke-Korsakoff syndrome (autopsies of 380 people with AIDS showed Wernicke’s encephalopathy in 10% of the cases)^65^, and these conditions are associated with thiamine deficiency. It is possible that patients with viral infection could have an increased risk of thiamine deficiency, but this information has remained largely unexplained in viral diseases^66^, including COVID-19. A potential reason could be that a deficiency in thiamine could be related to the thiamine transport protein, which can have a general preference for multiple membrane transport molecules which can function as receptors for candidate viruses^67^. The Th17 proinflammatory response has also been reported in the experimental encephalomyelitis model^57^. Thus, thiamine could be a therapeutic agent to alleviate neurological symptoms of COVID-19.

Adverse effects (AE) of thiamine are minimal and generally mild. Possible AEs include nausea, diarrhea, and abdominal pain. Rarely, individuals also suffer serious allergic reactions. There is no reported drug related symptoms at the 200 mg thiamine dose used in our study, and there are no reported AEs. A landmark pharmacokinetic study utilizing a 1500 mg maximum oral dose of thiamine in healthy subjects showed rapid absorption^32^. Moreover, 4000 mg thiamine administration showed no to mild AEs when used in children (Leigh’s disease). Thus, higher doses of thiamine for treatment of COVID-19’s cytokine storm could be considered a safe therapy.

Our study has several limitations. This is a small study. However, both clinical and *in vitro* evidence collectively support the potential of thiamine as a therapeutic agent in attenuating the Th17 proinflammatory response. We did not test the *in vitro/in vivo* efficacy of thiamine in the treatment of COVID-19 or its derivative stimulated Th17 proinflammatory response directly. We anticipate conducting such *in vitro* experiments for COVID-19 as a continuation of this project. Moreover, plasma thiamine levels have not been assessed in COVID-19. Our study did not have sufficiently large numbers of males and females; thus, identifying sex-differences was not within the scope of this study.

In summary, Thiamine has been approved by the Food and Drug Administration (FDA) of the USA as a prescription product and is considered very safe even at higher doses since it is water soluble and can be excreted via urine, if in excess^68^. Given its robust safety record, we suggest that thiamine should be considered for COVID-19 treatment studies.

## Data Availability

The datasets analyzed during the current study are available from the corresponding author by request.

## Acknowledgments

We thank research/clinical staff of the University of Louisville for their support. We thank Ms. Marion McClain for editing this manuscript.

## Ethics approval and consent to participate

Study was approved by the sites’ Institutional Review Boards (Ethics committees). All patients included in this study consented to participate before the beginning of the study.

## Disclosure

This article is not published nor is under consideration for publication elsewhere at the time of submission.

## Abbreviations

COVID-19: Coronavirus Disease-19
IL-17: Interleukin 17
IL-22: Interleukin 22
Disease Controls: (DC)
HV: Healthy Volunteers

## Trial registration

https://ClinicalTrials.gov identifier – NCT# 01809132 and 00106106.

## Reprint requests

Vatsalya Vatsalya MD, PgD, MSc, MS, Department of Medicine, University of Louisville School of Medicine, 505 S. Hancock St., CTR Room 514A, Louisville KY 40202 USA. Tel.: 502-852-8928. Fax: 502-852-8927.

## Notes

**Conflicts of Interest:** All authors declare no conflict of interests.

**Project/Grant Support**: Study was supported by NIH: NIH-OD-OHSRP-12176 (VV), R15CA170091-01A1 (MK), ZIA AA000466 (VAR), R01AA023190 (WF), P50AA024337-8301 and P20GM113226-6169 (XZ), and P50AA024337, P20GM113226, U01AA026936, U01AA0279880, R01AA023681 (CJM). The content is solely the responsibility of the authors and does not necessarily represent the official views of the National Institutes of Health.

### Competing Interest Statement

The authors have declared no competing interest.

### Clinical Trial

NCT# 01809132 and 00106106

### Funding Statement

Study was supported by NIH: NIH-OD-OHSRP-12176 (VV), R15CA170091-01A1 (MK), ZIA AA000466 (VAR), R01AA023190 (WF), P50AA024337-8301 and P20GM113226-6169 (XZ), and P50AA024337, P20GM113226, U01AA026936, U01AA0279880, R01AA023681 (CJM). The content is solely the responsibility of the authors and does not necessarily represent the official views of the National Institutes of Health.

### Author Declarations

Study was approved by the Institutional Review Boards of all the sites. All patients and healthy volunteers included in this study consented to participate before the beginning of the study.

